# Data-driven Subtyping and Staging of ALS: A Multicentre, Longitudinal, Deformation-Based Morphometry Study

**DOI:** 10.64898/2025.12.02.25341482

**Authors:** Isabelle Lajoie, Canadian ALS Neuroimaging Consortium (CALSNIC), Sanjay Kalra, Mahsa Dadar

**Author notes:** Correspondence to: Isabelle Lajoie, Mahsa Dadar, Cerebral Imaging Centre, 6875 Boulevard LaSalle, Montréal, QC, H4H 1R3. Co-senior authors (Sanjay Kalra, Mahsa Dadar).

## Abstract

Amyotrophic lateral sclerosis (ALS) is clinically and biologically heterogeneous, yet data-driven imaging subtyping approaches have rarely been validated longitudinally or linked to clinical and survival outcomes. We aimed to identify and validate distinct ALS subtypes and disease stages using deformation-based morphometry (DBM) and the Subtype and Stage Inference (SuStaIn) model, and to characterize their cross-sectional and longitudinal imaging, clinical, cognitive, and survival profiles.

Data from 198 ALS patients and 144 healthy controls in the CALSNIC multicentre cohort were analyzed. Baseline regional DBM w-scores from 14 ALS-relevant regions served as input to SuStaIn to infer subtypes and stages. Longitudinal consistency of subtype and stage assignments (e.g. adherence to the expected disease evolution) was assessed using follow-up visits. Imaging and clinical trajectories were compared across subtypes using linear mixed-effects models incorporating stage and elapsed time. Associations between longitudinal variables and SuStaIn stage were estimated using mixed models, while baseline clinical and cognitive differences were assessed with ordinary least squares regression. Survival differences were evaluated using Kaplan-Meier curves and log-rank tests.

SuStaIn identified one normal-appearing group (S0) and three ALS atrophy subtypes. S0 showed no baseline atrophy but exhibited longitudinal motor decline and the most favorable survival (log-rank *p* < 0.05 to *p* < 0.01). S1 exhibited classical motor/corticospinal tract-dominant degeneration, greater lower motor neuron burden, and intermediate survival. S2 showed limbic-onset atrophy progressing toward motor pathways, with preserved cognition and a milder course. S3 demonstrated extensive fronto-parietal and striatal atrophy, longitudinal motor-thalamic degeneration, and the shortest survival. Subtype and stage assignments demonstrated high longitudinal consistency (>90%). SuStaIn stage was strongly associated with widespread brain atrophy (and ventricular expansion), with the strongest effects in limbic-subcortical regions. Stage also correlated with ALSFRS-R decline and forced vital capacity reduction, indicating that stage reflects disease-linked progression.

This study establishes a robust, longitudinally validated model of ALS heterogeneity, showing that SuStaIn-derived subtypes define distinct disease trajectories, whereas the normal-appearing group reflects an early, structurally preserved state with a more favorable survival profile. By integrating probabilistic staging with longitudinal modelling, these findings clarify dynamic subtype-specific progression patterns and support the use of SuStaIn for biologically informed patient stratification, prognostication, and clinical trial enrichment in ALS.

## Introduction

Amyotrophic lateral sclerosis (ALS) is a fatal neurodegenerative disease characterized by progressive degeneration of both upper and lower motor neurons, leading to muscle weakness, respiratory failure, and death typically within 2-5 years of symptom onset.^1,2^ Clinical presentations are highly heterogeneous regarding site of onset (which may be bulbar, limb, or respiratory), progression rate, survival time, and cognitive/behavioral involvement.^3,4^ Thus, ALS lies within a spectrum (ALS-frontotemporal spectrum disorder [ALS-FTSD]); up to half of patients develop cognitive or behavioral impairment, and approximately 15% meet diagnostic criteria for the frontotemporal dementia.^5,6^

This heterogeneity complicates diagnosis, prognosis, and the interpretation of neuroimaging findings, as well as the design of therapeutic trials. Understanding subtype-specific trajectories allows for more precise intervention timing: rapid cognitive-motor decline may require early neuropsychological monitoring and multidisciplinary management, whereas “neuroprotected” individuals with slower progression could inform resilience mechanisms and targeted therapies.

Several clinical staging systems have been proposed to quantify ALS progression, most notably the King’s and Milano-Torino (MiToS) frameworks.^7,8^ These systems measure how a person’s ability to function declines over time, but they mainly focus on obvious motor symptoms and do not fully reflect the underlying biological changes in the disease. In contrast, post-mortem and neuroimaging studies suggest that ALS pathology spreads along distinct neuroanatomical pathways, starting primarily in motor regions such as the precentral gyrus and corticospinal tract, and progressing to extra-motor and limbic structures including the hippocampus and amygdala.^9–12^ Such complexity cannot be captured by linear clinical staging alone and highlights the need for data-driven, biomarker-based models that integrate both spatial and temporal dimensions of disease progression.

Magnetic resonance imaging (MRI) offers a sensitive in-vivo measure of ALS-related neurodegeneration. Deformation-based morphometry (DBM), in particular, provides a voxel-wise, whole-brain measurement of regional tissue deformation, quantifying both cortical and subcortical contraction (reflecting parenchymal atrophy) and expansion (such as ventricular enlargement).^13^ Previous DBM-based studies have detected progressive cerebral atrophy and expansion in various neurodegenerative disorders.^9,14–17^ However, conventional group-level approaches typically assume a single progression pattern across patients, thereby obscuring meaningful biological variability. Recent advances in machine learning have enabled the development of probabilistic disease progression models capable of revealing hidden subtypes and their temporal evolution within a population.^18–20^

The Subtype and Stage Inference (SuStaIn) model is one such framework. SuStaIn jointly infers distinct disease subtypes, each representing a characteristic sequence of biomarker abnormalities, and assigns every participant a probabilistic stage along that trajectory.^18^ Originally developed for Alzheimer’s disease,^18,21^ SuStaIn has since been applied to frontotemporal dementia,^22^ multiple sclerosis,^23^ and more recently to TDP-43 pathology and ALS-FTD.^24,25^ Yet, prior ALS applications were limited by primarily cross-sectional data, and limited validation against longitudinal clinical or imaging outcomes. Moreover, the relationship between SuStaIn-derived disease stage and both clinical and imaging measures remains largely unexplored. Notably, no prior studies have leveraged DBM within a multicenter, longitudinal framework to capture subtype-specific trajectories and evaluate the temporal consistency of SuStaIn-derived stages.

The present study addresses these limitations by applying SuStaIn to DBM-derived regional brain volumes from a large, prospectively acquired multicenter longitudinal ALS cohort (CALSNIC-1 and CALSNIC-2). Our goals were to [i] identify data-driven ALS subtypes and their progression trajectories; [ii] evaluate the longitudinal consistency of subtype and stage assignments; [iii] compare cross-sectional and longitudinal imaging and clinical features between subtypes, including survival; and [iv] examine the association between inferred disease stage and both clinical and imaging measures, accounting for elapsed time. By integrating SuStaIn with longitudinal mixed-effects modeling, this work aims to define biologically meaningful trajectories of ALS progression that integrate imaging-based staging with clinical outcomes.

## Materials and methods

### Participants

We used longitudinal (baseline, visit2 ∼ month 4, visit3 ∼ month 8) MRI and clinical measurements of ALS patients and healthy controls (HCs) from the CALSNIC study.^26^ CALSNIC adheres to the principles of the Declaration of Helsinki and received approval from the Health Research Ethics Boards of all participating sites. Written informed consent was obtained from all participants and the University of Alberta Health Research Ethics Board (HREB) granted approval for the protocol presented in this study. Clinical and MRI protocols were harmonized across research centers and vendors. Patients were included if they had signs of both UMN and LMN involvement upon initial diagnosis and met criteria for possible, probable, laboratory-supported probable, or definite ALS according to the Revised El Escorial Criteria.^27^ Following the exclusion criteria (detailed in the Supplementary Materials, Section A), the final study population included 198 ALS patients (198_baseline_/120_visit2_/80_visit3_) and 144 HCs (144_baseline_/110_visit2_/84_visit3_).

### Clinical and Demographic Data

Clinical evaluation included the ALS Functional Rating Scale-Revised (ALSFRS-R),^28^ disease progression rate (DPR), finger and foot tapping rates, Edinburgh Cognitive and Behavioural ALS Screen (ECAS),^29^ cognitive impairment^30^ transformed into percentage of abnormal values, UMN^26^ and LMN involvement, site of (first symptom) onset, forced vital capacity (FVC), symptom duration at the first MRI visit, years of education, age and sex. For details on the clinical evaluations, see Supplementary Materials Section B.

### Image Preprocessing

All T1-weighted MRIs were processed using PELICAN, our open source and extensively validated pipeline (https://github.com/VANDAlab/Preprocessing_Pipeline) based on the open access MINC (https://github.com/BIC-MNI/minc-tools) and ANTs (http://stnava.github.io/ANTs/) tools.^31^ Pre-processing steps included: denoising,^32^ intensity inhomogeneity correction,^33^ and intensity normalization using histogram matching. The images were linearly^34^ and nonlinearly^35^ registered to the MNI-ICBM152-2009c template.^36^ All steps were visually assessed to exclude cases with significant artifacts or inaccurate registrations. DBM maps were calculated as the Jacobian determinant of the non-linear deformation fields. DBM values greater than one indicates localized expansion compared to the corresponding voxels in the template, whereas values smaller than one indicate relative shrinkage, i.e. atrophy. For regional analyses, DBM maps were masked to remove the partial volume effects of the sulci and mean DBM was calculated based on 62 bilateral GM and ventricle regions from the CerebrA atlas,^37^ and WM tracts and corpus callosum based on the JHU^38^ and Allen^39^ atlases, respectively.

To focus on disease-specific atrophy in ALS patients, we calculated regional w-scores at each patient visit.^40^ The w-score is the adjusted difference between a patient’s observed DBM value and its predicted value normalized for age, sex, and imaging site. This predicted value is derived from a linear mixed-effects model based on data from HCs. A negative w-score indicates greater atrophy compared to expected levels in the HCs.

### Subtype and Stage Inference Modelling

The Subtype and Stage Inference (SuStaIn) algorithm is an unsupervised machine-learning framework that jointly models disease subtypes and within-subtype progression stages from cross-sectional data.^18^ SuStaIn identifies subgroups of individuals sharing similar patterns of biomarker abnormality and reconstructs a probabilistic sequence of disease progression within each subtype. Each participant is then assigned both a most likely subtype and an estimated disease stage.

SuStaIn models disease progression as a sequence of biomarkers transitioning from normal to abnormal states, relative to a control group. In this study, biomarkers corresponded to regional brain volumes derived from DBM-based w-scores, interpreted as measures of atrophy. The algorithm thus infers distinct spatiotemporal trajectories of regional volume loss, reflecting heterogeneous patterns of ALS neurodegeneration. The present analysis employed the Python implementation of SuStaIn.^41^

Given SuStaIn’s modeling constraints favoring parsimony in biomarker number relative to sample size, we selected 14 regions combining core loci from the canonical Brettschneider pTDP-43 staging scheme^12^ with additional areas like superior parietal, well supported in neuroimaging ALS progression literature.^42,43^ Our features capture fundamental motor/premotor cortices (precentral, paracentral), key white matter tracts (corticospinal tract, brainstem), association cortices (superior frontal, rostral middle frontal, inferior and superior parietal), pivotal subcortical nuclei (caudate, putamen, amygdala), and medial temporal structures (hippocampus, entorhinal, parahippocampal cortex). This set balances biological specificity and sensitivity, reflecting both histopathology and neuroimaging, and strengthens interpretability and statistical power in subtype inference. For each ROI, two w-score abnormality thresholds (1 and 2) were used, yielding 28 model events. The maximum w-score threshold per region was set at 2 or 3 depending on the regional z-score distribution.

Model fitting was performed on baseline data, with 8-fold cross-validation used to determine the optimal number of subtypes (*k*). Model selection was guided by the Cross-Validation Information Criterion (CVIC), which balances model fit and complexity (lower values indicate better fit), and the out-of-sample log-likelihood, assessing predictive generalization (higher values indicate better fit). Model uncertainty was estimated through 10,000 Markov Chain Monte Carlo (MCMC) iterations.

A single-subtype SuStaIn model (*k*=1) was fitted using the same input parameters to provide a continuous, unified stage estimate across all patients. This cohort-level progression trajectory was primarily used to benchmark the model’s overall staging sequence against the established Brettschneider pathological stages.^12^

Participant-level subtype and stage assignments were obtained by maximizing the posterior likelihood of stage given the participant’s regional biomarker profile within each subtype. Individuals assigned to stage 0, indicating no abnormal regional w-scores, were classified as Normal Appearing (S0), while those with stage > 0 were assigned to their most probable subtype.^18^

### Longitudinal Consistency

The trained SuStaIn model derived from baseline data was subsequently applied to the longitudinal MRI scans to infer subtype and stage assignments at each available follow-up visit. This allowed assessment of the temporal consistency of the model’s subtype and stage predictions within individuals over time.

Subtype assignment was defined as *consistent* if participants either (i) remained assigned to the same subtype across visits or (ii) progressed from the Normal Appearing group (stage 0, i.e., no detectable atrophy at baseline) to any SuStaIn-defined subtype. Conversely, changes from one established subtype to another (e.g., S1 → S3) were considered *inconsistent*.

Stage evolution was defined as *consistent* if a participant remained at the same stage or progressed to a later stage at follow-up, reflecting monotonic disease advancement. Regression to an earlier stage was classified as *inconsistent*.

To evaluate assignment confidence, subtype and stage probabilities at the earliest visit were compared between *consistent* and *inconsistent* cases using a Student’s t-test for normally distributed data or a Mann-Whitney U test otherwise.

To ensure consistent and interpretable subtype characterization for group-level analyses, patients exhibiting longitudinal subtype transitions were assigned a single, most confident subtype. Specifically, for individuals whose inferred subtype changed across visits, the subtype with the highest posterior assignment probability across all time points, excluding S0 (Normal Appearing), was selected as their final representative subtype. Importantly, disease stage values were retained across visits to accurately reflect their individual disease progression timeline. For example, a participant moving from S0 to S1 would retain their previous stage (stage=0), thereby maintaining information about the individual’s prior S0 status while allowing for a coherent subtype classification. This approach maintains the temporal integrity of SuStaIn-derived stages while ensuring that each participant contributes a single, most probable subtype to downstream clinical and imaging analyses, thereby improving model consistency and biological interpretability.

### Statistical Analyses

#### Cross-sectional and Longitudinal Group Comparisons

Group comparisons of longitudinal continuous clinical features were performed using a linear mixed-effects (LME) model applied to the longitudinal data, specified as follows:

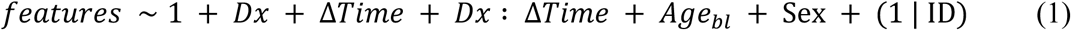

where Dx is a categorical fixed effect contrasting groups (ALS vs controls, or SuStaIn-inferred ALS subtypes) to assess group-level and longitudinal differences. The Dx:ΔTime interaction term captures group differences in the rate (slope) of longitudinal change. Age_bl_ represents age at baseline (first visit), and Sex is a categorical covariate contrasting males and females. Participant ID was included as a random intercept to account for within-subject correlations across repeated measures.

For comparisons among ALS subtypes, the model additionally controlled for the confounding effect of disease progression (i.e., differences in baseline atrophy levels) by including the inferred disease stage at baseline (Stage_bl_) as a covariate. This ensures we isolate regional atrophy differences that are uniquely linked to the subtypes themselves.

Group differences in baseline demographic and clinical variables (e.g., age, sex, years of education, site of onset, ECAS, ALSFRS-R, DPR) were assessed using regression models appropriate to variable type. Continuous variables were analyzed using ordinary least squares (OLS) regression, binary variables using logistic regression, and ordered categorical variables using an ordinal logistic regression model.

For imaging analyses, regional w-scores were compared using an analogous LME model with the following specification:

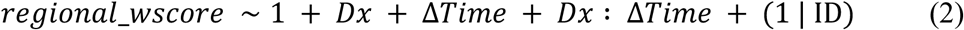

In this case, Age_bl_, Sex, and Scanner effects had already been accounted for during the computation of the w-scores. The same modeling framework was applied for comparisons among ALS subtypes, including Stage_bl_ as an additional covariate.

For each set of group comparisons (e.g., S1 vs. S2, ALS vs. HCs), correction for multiple comparisons was performed by controlling the False Discovery Rate (FDR) at a level of *p* < 0.05. This correction was applied independently across two distinct sets of features: one for all clinical measures, and one for all imaging measures.

#### Survival Analysis

Survival outcome was defined as the composite endpoint of death, tracheostomy, or assisted ventilation for a minimum of 22 hours per day. Patients were censored if they were alive five years after the baseline MRI visit, lost to follow-up, or if they opted for medical assistance in dying (MAID). The final survival analysis cohort included 185 out of 198 patients, all of whom had follow-up data after the initial MRI visit, resulting in a censoring rate of 56%.

A Kaplan-Meier (KM) model was employed to visually represent the survival probabilities over time for each of the inferred ALS subtypes. The statistical significance of the difference in survival distributions between these subtypes was formally determined using the log-rank test.

#### Association Analysis: Clinical and Imaging Features with Disease Stages

In addition to conventional longitudinal modeling of atrophy progression over time (Dx:Δtime), we performed association analyses between SusTaIn-inferred disease stage and both imaging and clinical features. While longitudinal LME models capture linear, time-dependent change, the stage-based approach evaluates disease-linked progression, that is, how strongly each feature co-varies with the probabilistic disease stage inferred by the SuStaIn model. By including ΔTime as a covariate, this analysis isolates disease-related variance from simple effects of elapsed time, allowing us to identify brain regions and clinical measures most tightly coupled to disease advancement along each subtype’s inferred trajectory. Together, the stage- and time-based analyses provide complementary perspectives on ALS progression and serve as a validation of the biological and clinical relevance of the SuStaIn staging model.

The association between features and disease stages was assessed within each subtype, and subsequently across all ALS patients combined (including the S0 group). Baseline demographic and clinical variables were analyzed using OLS regression, with Stage as the primary predictor. Longitudinal clinical features were assessed using a LME model with Stage as the primary predictor, ΔTime as a fixed effect to account for the natural and linear effect of elapsed time, and participant ID as a random intercept to account for repeated measures. For both baseline and longitudinal clinical analyses, Age_bl_ and Sex were included as covariates when appropriate, with years of education additionally included for models involving the continuous raw ECAS cognitive metrics.

Similarly, the association with longitudinal imaging features (w-scores) was also assessed using an LME model with Stage and ΔTime as fixed effect and ID as a random intercept. Notably, Age_bl_ and Sex were omitted from the imaging LME model, as their effects had already been accounted for during the w-score normalization process.

Correction for multiple comparisons was applied to the entire set of association tests. This was achieved by controlling the FDR across all calculated *p*-values (for both clinical and imaging associations) at a level of *p* < 0.05.

## Results

### Cohort Characteristics

One hundred forty-four HCs and 198 patients with ALS were included in this study. A summary of the participants’ demographics, cognitive, and functional performance is presented in Table 1. Mean age in the patients with ALS was significantly greater than that of HCs. There was a greater proportion of men in the patients with ALS than the HCs. Years of education were slightly lower in the patients with ALS compared with the HCs. As expected, the patients with ALS showed greater cognitive and functional impairment. Significant differences were found in finger and foot tapping, and ECAS scores.

**Table 1.** Baseline Cohorts Demographics.

|  | **HCs** | **Patients With ALS** | **p value** |
| --- | --- | --- | --- |
| **N visit (v1/v2/v3)** | 144/110/84 | 198/120/80 | 0.16<sup>a</sup> |
| **Age, yr** | 57 (40-77) | 60 (40-86) | <0.01<sup>b</sup> |
| **Sex, M:F** | 62:82 | 128:70 | <0.01<sup>b</sup> |
| **Years Education, yr** | 16 (7-28) | 15 (4-28) | <0.05<sup>b</sup> |
| **Finger Tapping** | 57 (26-90) | 41 (0-83) | <0.0001<sup>c</sup> |
| **Foot Tapping** | 41 (18-76) | 24 (0-64) | <0.0001<sup>c</sup> |
| **ECAS Total Score** | 116 (91-134) | 108 (47-127) | <0.0001<sup>b</sup> |

Values are expressed as median (range). Statistical significance is indicated in bold ( $p < 0.05$ ). Group comparisons were performed using:
<sup>a</sup> Ordinal logistic regression for ordered categorical variables.
<sup>b</sup> Ordinary least squares (OLS) regression for baseline continuous variables.
<sup>c</sup> Linear mixed-effects (LME) models for longitudinal continuous variables.
ALS = amyotrophic lateral sclerosis; ECAS = Edinburgh Cognitive and Behavioural ALS Screen.

### SuStaIn Input Features and Optimal Number of Subtypes

Fig. 1A shows the SuStaIn-derived sequence of regional progression when restricting to a single ALS subtype. The w-score heatmap highlights early, pronounced involvement of the primary motor cortex (precentral, paracentral), corticospinal tract, and brainstem, which is consistent with Brettschneider’s stage 1 pathological spread. This is further illustrated in Fig. 1B (top row), where SuStaIn stage 3 predominantly displays motor cortex and brainstem atrophy. As progression advances, the distribution expands to include parietal regions (stage 4), followed by medial temporal structures: amygdala, hippocampus, entorhinal, and parahippocampal cortices (between stages 4 and 13). Subcortical nuclei (caudate, putamen) emerge between stages 8 and 15, with association cortices (superior frontal, rostral middle frontal) affected at later stages (>13).

**Figure 1.**
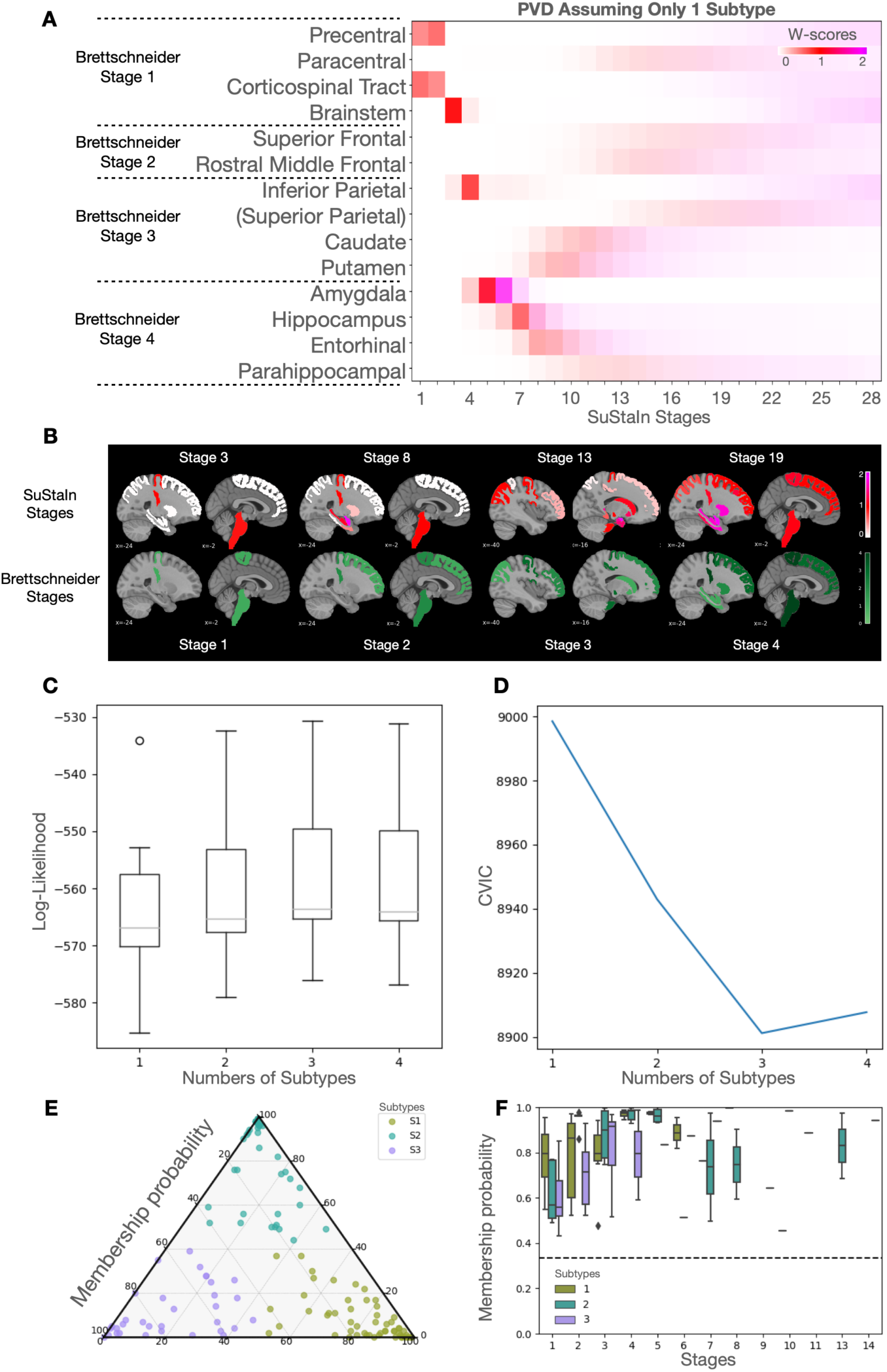
Subtype and stage inference modeling. **(A)** displays the Positional Variance Diagrams (PVDs) obtained when the SusTaIn model was initially run assuming only one subtype. This PVD illustrates the single, assumed sequence of regional atrophy progression, with the X-axis representing the SusTaIn disease stage (1-28) and the Y-axis listing Regions of Interest (ROIs). Color intensity reflects the severity threshold reached (w-score=1 in red; =2 in magenta) and the model’s certainty (darker color = high certainty). **(B) (Top)** shows the spatial distribution of brain atrophy patterns across selected SuStaIn stages (3, 8, 13, 19), with the color bar indicating the cumulative probability of abnormality. **(Bottom)** depicts the corresponding canonical Brettschneider pathological stages (1-4) mapped anatomically for comparison. The color bar indicates the cumulative count of stages since a region has appeared. **(C)** and **(D)** show, for each subtype model (k=1-4), the distribution of average negative log-likelihood **(C)**, and the Cross-Validation Information Criterion (CVIC) **(D)**, respectively. Higher log-likelihood and lower CVIC represents better model fit. **(E)** is a Ternary Plot that shows the membership probability for individual patients (dots) across the three inferred subtypes (S1, S2, S3). **(F)** displays the membership probability of assigned subtypes across the inferred disease stages. The dotted line represents the chance level (random assignment probability), which is confidently exceeded across all non-zero stages.

The bottom row in Fig. 1B depicts the corresponding Brettschneider stages, mapped to canonical anatomical blocks: stage 1 aligns well with early motor and brainstem involvement; stage 2 localizes to superior and middle frontal regions; stage 3 encompasses parietal cortex and striatal nuclei; and stage 4 includes limbic temporal regions. When directly compared, the SuStaIn model recapitulates the major axis of ALS pathological progression described by Brettschneider but shows delayed involvement of prefrontal and striatal regions. These differences may reflect cohort variability and the sensitivity of MRI-based progression modeling to extra-motor and association cortical involvement.

Fig. 1C and 1D show the test set log-likelihood and the Cross Validation Information Criterion (CVIC) respectively for determining the optimal number of ALS subtypes (*k*). An optimum is clearly obtained at *k*=3 subtypes, indicated by a log-likelihood that stabilizes after increasing as *k* increases from 1 to 3, and the CVIC reaching its minimum value. Fig. 1E illustrates the three subtypes membership probability for each patient, all stages combined, while 1.F shows the boxplots representing subtype membership probability according to the inferred disease stage. The subtype assignment probability remained significantly above the chance threshold (indicated by the dotted line) throughout all non-zero SuStaIn stages, demonstrating the model’s high confidence in its classification.

### SuStaIn-Inferred Trajectories

Fig. 2 illustrates the spatiotemporal progression of atrophy estimated by SuStaIn across the three ALS subtypes. Each panel shows the sequence in which regional deformation (W-score) abnormalities appear and spread across disease stages. Subtype 1 shows a motor-predominant pattern consistent with classical ALS progression. Early stages (Stages 1-6) reveal initial atrophy in the CST, followed by the brainstem, precentral, and paracentral gyri. As disease stages advance (from Stages 7), atrophy extends to frontoparietal and striatal structures (putamen), and finally involves limbic and medial temporal regions (amygdala, hippocampus, entorhinal, parahippocampal cortex) at later stages (after stage 10), terminating with an atrophy in caudate, from stage 16. Subtype 2 displays a limbic-onset pattern. The earliest stages (Stages 1-7) are characterized by marked atrophy in amygdala, entorhinal cortex, and hippocampus, indicating early limbic involvement. Atrophy of the CST emerges around Stage 5, followed by progressive extension to precentral gyrus, brainstem, inferior parietal, caudate, putamen, and parahippocampal regions. Later stages (after Stage 16) show additional frontal and parietal cortical involvement (superior frontal, rostral middle frontal, superior parietal), suggesting a spread from limbic to motor and associative cortices. Subtype 3 demonstrates a motor-frontostriatal pattern. The first affected regions include the precentral gyrus, followed by superior and inferior frontal and inferior parietal cortices, together with caudate at Stage 2. Progression continues with putaminal and amygdalar involvement, later reaching paracentral areas and ultimately the brainstem in advanced stages.

**Figure 2.**
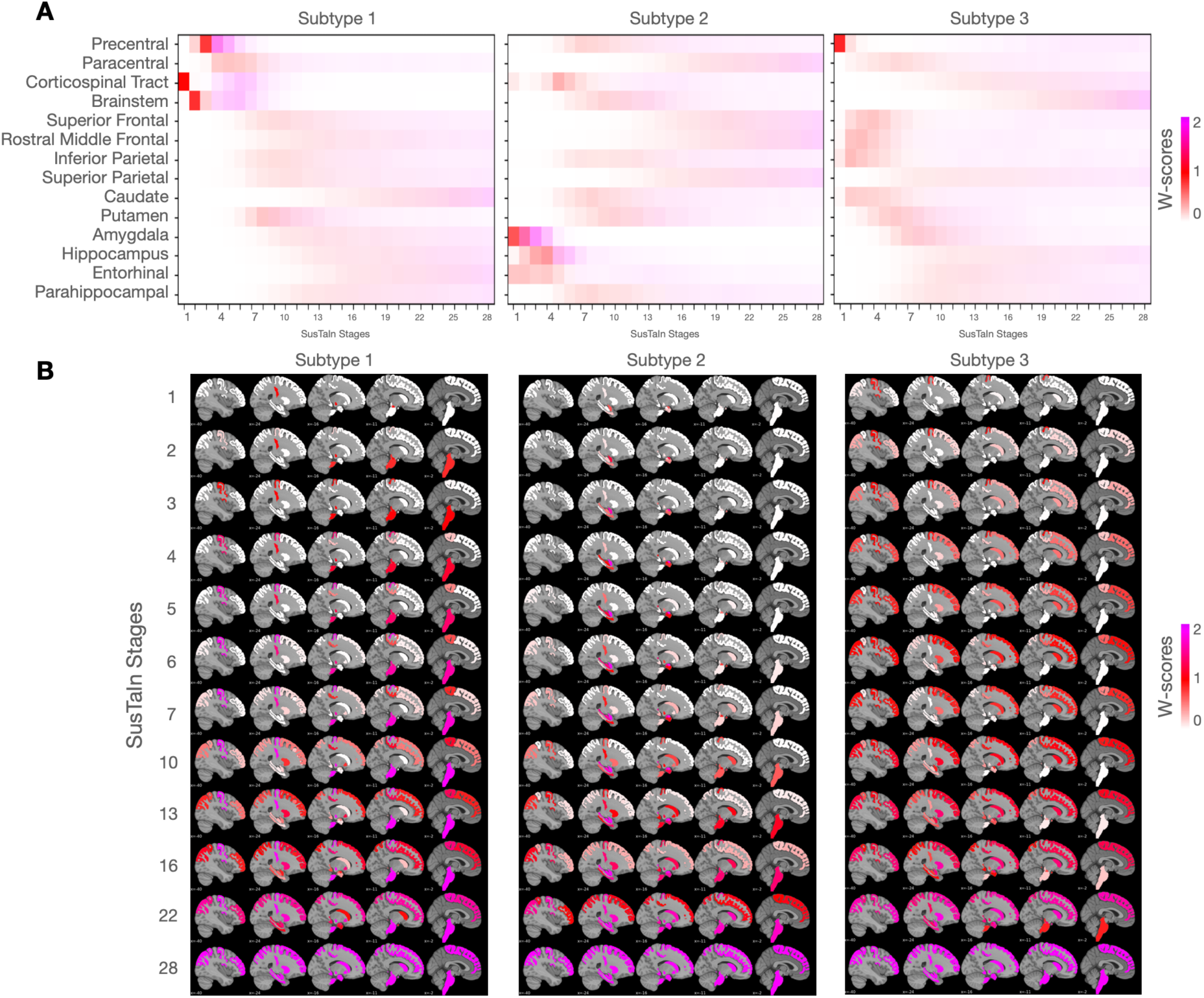
Progression patterns. Inferred trajectory of regional atrophy progression in subtypes. **(A)** displays the Positional Variance Diagrams (PVDs), representing the inferred sequence and variability of regional atrophy progression for each subtype. The X-axis is the derived disease stage, while the Y-axis lists the specific ROIs. Each colored block indicates the stage at which an ROI crosses a pathological threshold, with the color corresponding to the severity score threshold reached (w-score =1 in red; =2 in magenta). The density of the color reflects the model certainty regarding that region’s position in the overall sequence, where darker colors indicate high certainty (low positional variance) and lighter colors indicate lower certainty. **(B)** presents the corresponding regional cumulative probability of abnormality overlaying brain sagittal views, illustrating the overall spatial pattern of atrophy at different stages of the inferred trajectory for each subtype.

### Longitudinal Consistency

Fig. 3 illustrates the longitudinal consistency of SuStaIn subtype and stage assignments across MRI visits using Sankey diagrams (top) and corresponding box plots of assignment probabilities (bottom). Subtype consistency (Panel A) was high overall, with 92% consistency between V1-V2, 95% between V2-V3, and 91% between V1-V3. Most participants either remained within the same subtype across visits or transitioned from S0 to a subtype, both patterns being considered consistent with expected disease evolution. This pattern indicates robust reproducibility of SuStaIn subtype classification over time. In contrast, cases that transitioned between subtypes (“inconsistent”) showed significantly lower subtype assignment probabilities than consistent cases across all intervals (*p* < 0.05 to *p* < 0.0001), suggesting that misclassification primarily occurred among participants with less confident baseline assignments. Stage consistency (Panel B) was even higher, reaching 97% consistency from V1-V2, 92% from V2-V3, and 96% from V1-V3. Similar to subtypes, inconsistent stage transitions (e.g., to a lower stage) were associated with lower assignment probabilities, although significance was reached only for the V2-V3 comparison (*p* < 0.001), with the remaining contrasts showing a non-significant trend toward lower probabilities in inconsistent cases.

**Figure 3.**
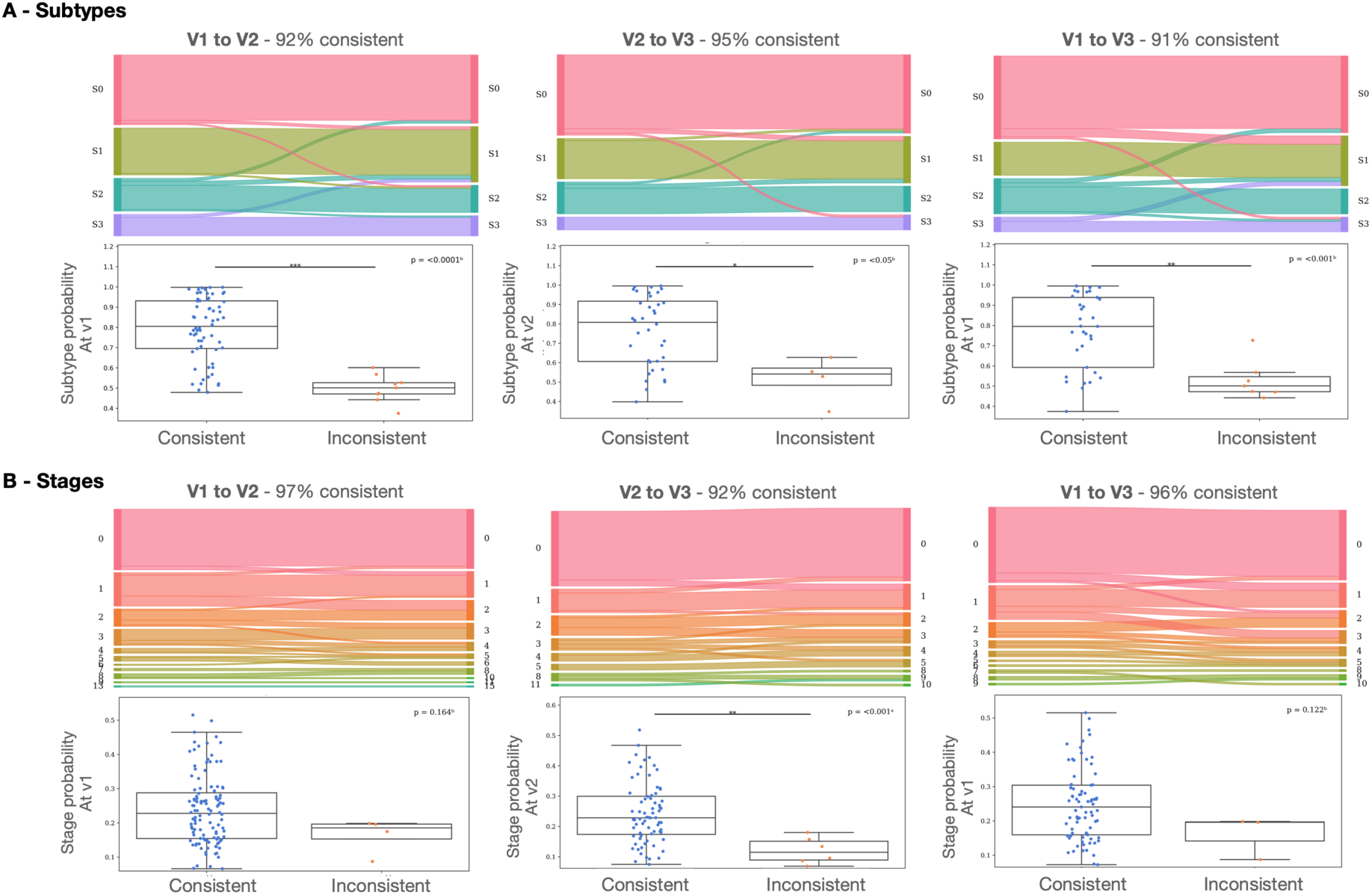
Longitudinal consistency of SuStaIn subtype and stage assignments. The Normal Appearing (S0) group was included, representing individuals assigned to Stage 0. Sankey diagrams **(Top)** and probability box plots **(Bottom)** illustrate the between-visit consistency of SuStaIn-inferred subtypes **(A)** and stages **(B)** across MRI timepoints: V1→V2 **(left)**, V2→V3 **(middle)**, and V1→V3 **(right)**. The Sankey flows are color-coded to represent the proportion of patients within each subtype or stage and their transitions across visits. Subtype and stage assignment probabilities from the earliest visit were compared between consistent and inconsistent cases using Student’s t-tests for normally distributed data or Mann-Whitney U tests otherwise. Statistical significance is denoted as follows: * *p* < 0.05, ** *p* < 0.01, *** *p* < 0.001. Consistent cases were defined as those showing (i) no subtype or stage change, (ii) a transition from S0 to a subtype, or (iii) progression to a higher disease stage.

Among patients with longitudinal data, 11 out of 198 showed a transition in their inferred subtype between visits. Accordingly, their most confident subtype (i.e., the subtype with the highest assignment probability across visits) was reassigned for subsequent clinical and imaging analyses. The transitions were distributed as follows: 5 from S0 to S1, 1 from S0 to S2, 1 from S0 to S3, 1 from S2 to S1, 1 from S2 to S3, and 2 from S3 to S1. This resulted in a final sample comprising 61 Normal Appearing (S0) individuals and 137 participants assigned to one of three SuStaIn-inferred ALS subtypes (S1 = 65, S2 = 39, S3 = 33).

### Demographic and Clinical Subtypes Comparisons

Analysis of baseline demographics and clinical characteristics (Fig. 4) revealed distinct profiles across the SuStaIn-derived subtypes. While most subgroups displayed comparable symptom duration, DPR, and education levels, S3 patients were the youngest cohort, exhibiting the lowest median age at the first visit (56 years old) and median age at onset (54 years old), although these differences did not reach statistical significance compared to others. FVC appeared higher in S0 compared to the subtype’s groups, but this difference also did not survive FDR correction. A major difference was observed in sex distribution: S2 exhibited a near-equal male-to-female ratio (17:22), suggesting a distinct female predominance compared to the predominantly male S0 (45:16), S1 (44:21), and S3 (22:11) cohorts. The El Escorial diagnosis also showed variation: S0 and S1 were most frequently diagnosed as “Probable”, while S3 was most frequently “Definite”. Notably, S2 had a high proportion of “Possible” and “Probable” diagnoses. All groups primarily presented with limb-only onset, with S0 showing the most substantial difference (53 limb-only vs 8 including bulbar). Finally, longitudinal attrition varied, with S1 maintaining the most comprehensive follow-up data (less than 30% attrition across three visits), whereas S3 suffered the highest percentage of attrition (over 50% loss between each visit), suggesting rapid progression or high burden impacting clinic attendance. None of the differences in features across subtypes were statistically significant.

**Figure 4.**
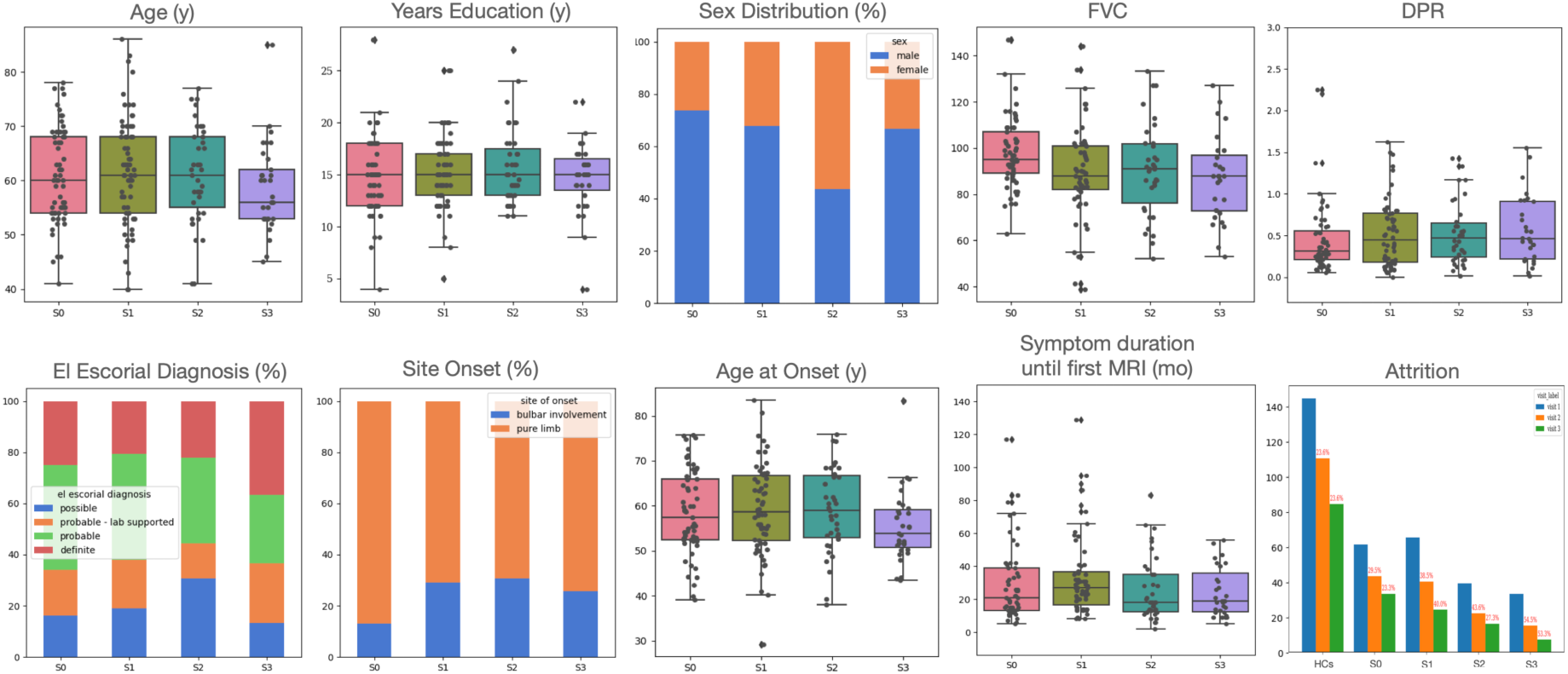
Baseline demographic and clinical features across SusTaIn-inferred ALS subtypes. The Normal Appearing (S0) group was included, representing individuals assigned to Stage 0. Ten comparative panels illustrate differences in age, education, sex, FVC, DPR, El Escorial diagnostic categories, site of onset, age at onset, symptom duration prior to MRI, and patient attrition among inferred ALS subtypes and S0 group. Box plots show medians, interquartile ranges, and outliers for continuous variables (age, education, FVC, DPR, age at onset, symptom duration); bar or stacked charts represent categorical variables (sex, El Escorial diagnosis, site of onset, attrition). Attrition panels indicate the number of patients per visit for each subtype and percentage lost per group in red (attrition includes censored and deceased patients). Group differences for continuous baseline variables were analyzed using OLS regression, while repeated longitudinal measures were assessed using LME models. Binary outcomes were evaluated via logistic regression and ordered categorical outcomes by ordinal logistic regression. Multiple comparisons were FDR-corrected (*p*<0.05). No differences between subtypes were statistically significant after correction. ALS = Amyotrophic Lateral Sclerosis; DPR = Disease Progression Rate, estimated using (48-ALSFRS-R)/symptom duration; FDR = False Discovery Rate; FVC = Forced Vital Capacity; HCs = Healthy Controls; LME = Linear Mixed-Effects; MRI = Magnetic Resonance Imaging; OLS = Ordinary Least Squares.

Fig. 5 illustrates differences in clinical and cognitive function among subtypes. ECAS scores (Fig. 5A top) revealed distinct cognitive profiles, with varying degrees of impairment across domains and subtypes. S0 and S3 are associated with the highest rates of abnormal scores, indicating pronounced multidomain dysfunction. Specifically, S3 shows the greatest impairment in memory, language, and fluency domains, while S0 exhibits deficits in language and fluency. In contrast, S1 generally manifests only mild or minimal abnormalities, and S2 stands out with the lowest levels of cognitive dysfunction across all measured domains. Although rates of abnormal scores differ by subtype, these between-ALS subtype differences did not reach statistical significance within the cohort. Instead, significant differences in cognitive impairment were consistently identified only when comparing certain ALS subtypes to healthy controls, and only in specific ECAS domains. Radar plots (Fig. 5A bottom) further illustrate these patterns, showing that subtype 3 encompasses more widespread and severe cognitive compromise, followed by subtype 0, whereas subtype 2 remains the most spared.

**Figure 5.**
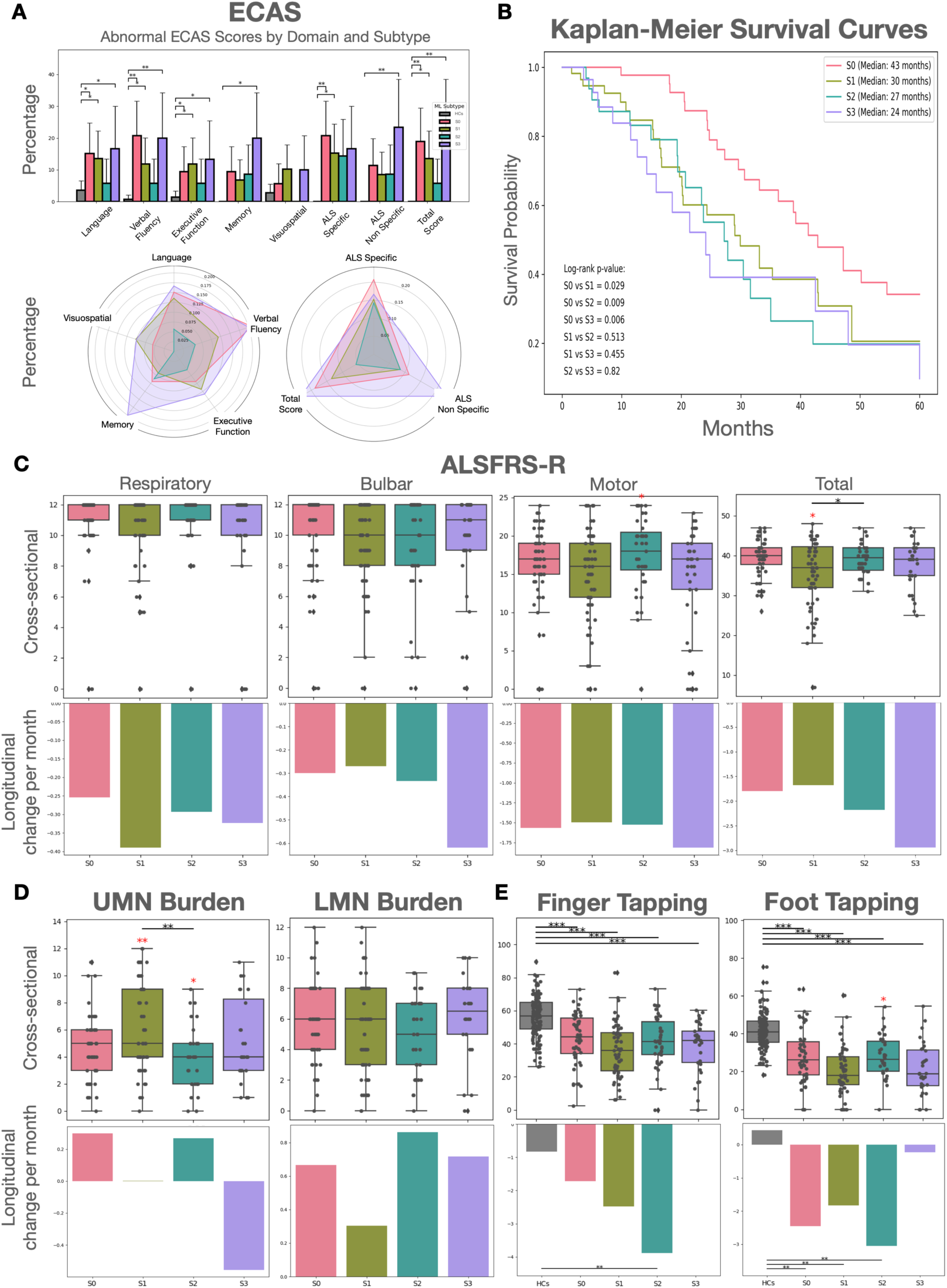
Cognitive, functional, motor, and survival differences across ALS subtypes. The Normal Appearing (S0) group was included, representing individuals assigned to Stage 0. Healthy controls (HCs) were also included when appropriate. **(A)** Bar graphs and radar plots illustrate abnormal ECAS scores by cognitive domain for ALS subtypes and HCs, including ALS specific (language, fluency, executive) and ALS non-specific (memory, visuospatial). **(B)** Kaplan-Meier survival curves compare probabilities and median survival durations for each subtype, with log-rank p-values indicated. **(C)** ALSFRS-R boxplots show cross-sectional respiratory, bulbar, motor, and total scores, while corresponding bar charts report each subtype’s mean longitudinal change per four months. **(D)** UMN and LMN burden scores are displayed using boxplots and mean longitudinal decline per four months bar charts per ALS subtype. **(E)** Finger and foot tapping performance for ALS subtypes and healthy controls are summarized by boxplots (cross-sectional) and bar charts (mean longitudinal change per four months). Significant group differences, with FDR correction at *p* < 0.05, are indicated by asterisks. Red asterisks denote a subtype’s difference compared to all other ALS subtypes combined. Statistical significance levels are represented as follows: * *p* < 0.05, ** *p* < 0.01, *** *p* < 0.001. Boxplots display the median, interquartile range, and outlying values for each group. Bar charts depict the mean monthly change per group. Group differences for repeated longitudinal measures were assessed using LME models. Binary outcomes were evaluated via logistic regression, and log-rank tests for survival data. ALS = amyotrophic lateral sclerosis; ALSFRS-R = Amyotrophic Lateral Sclerosis Functional Rating Scale-Revised; ECAS = Edinburgh Cognitive and Behavioural ALS Screen; FDR = False Discovery Rate; LME = Linear Mixed-Effects; LMN = Lower Motor Neuron; OLS = Ordinary Least Squares; UMN = Upper Motor Neuron.

The Kaplan-Meier Survival Curves (Fig. 5B) revealed the worst prognosis for the ALS subtypes: S1 (Median: 30 months) and S2 (Median: 27 months), with S3 exhibiting the shortest median survival time of 24 months. This was significantly lower than in the S0 group, which had a median survival of 43 months. Analysis of the ALSFRS-R Total score (Fig. 5C) showed S2 with the highest motor score when compared to all the other ALS patients, while S1 had the lowest ALSFRS Total score. Longitudinal analysis of the average change per month in ALSFRS-R scores did not yield significant differences between the cohorts. In terms of clinical disease burden (Fig. 5D), S1 carried a significantly higher cross-sectional UMN Burden score than all other patients, while S2 had the lowest. This was reflected in functional tests (Fig. 5E): S2 consistently exhibited the highest cross-sectional scores for both Finger Tapping (not significantly different) and Foot Tapping, indicating the best-preserved distal limb function. Longitudinally, S2 showed a pronounced decrease in Finger Tapping score compared to HCs and S3. Intriguingly, S3 was the only subtype whose Tapping score showed almost no longitudinal change (compared to that of controls).

### Cross-sectional contrasts confirmed separable imaging signatures

Cross-sectional contrasts against healthy controls (Fig. 6A, left) revealed four separable imaging signatures.

**Figure 6.**
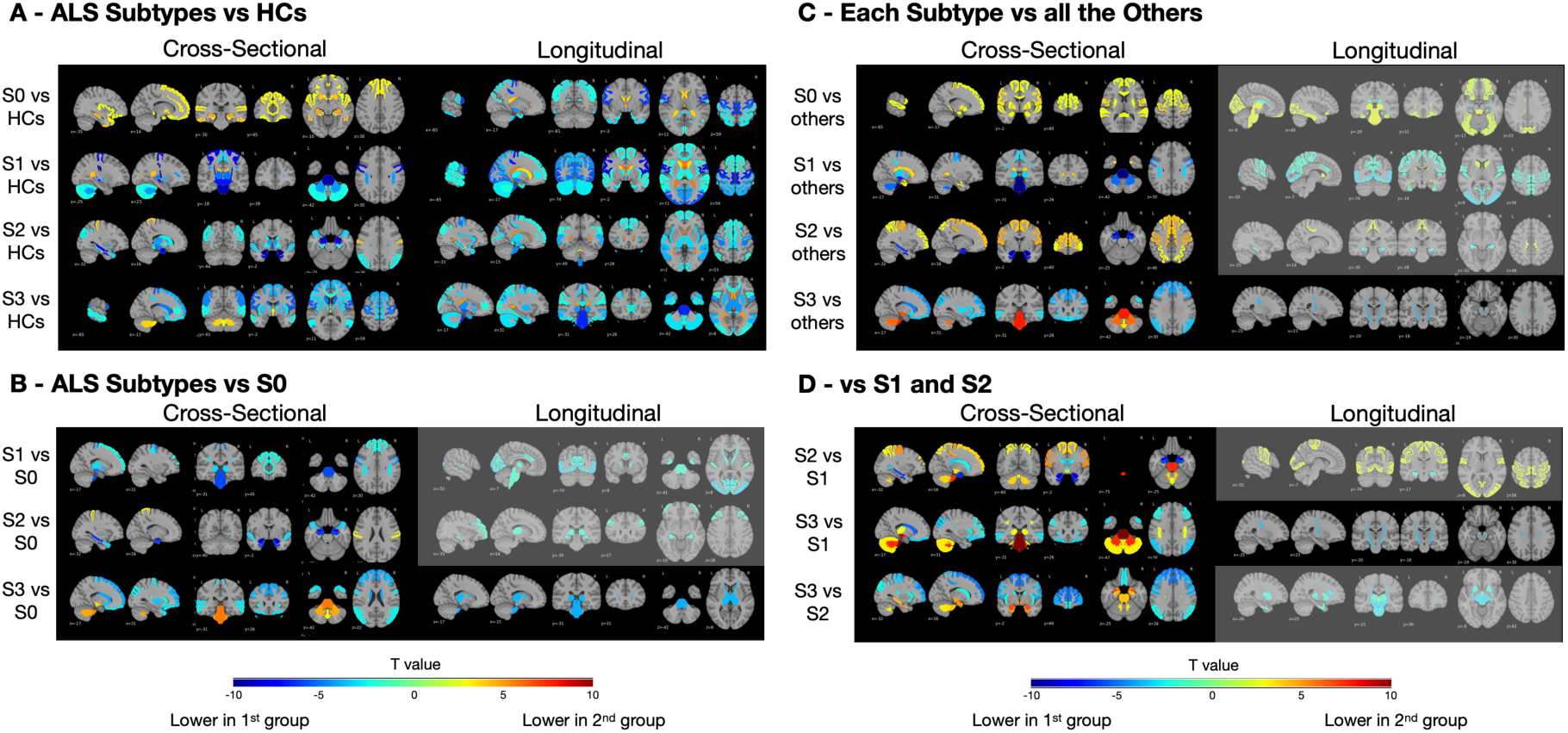
Maps of brain volume differences across SusTaIn-inferred subtypes, S0 and HCs using regional DBM w-scores. **(A)** Cross-sectional and longitudinal contrasts of S0 and each SusTaIn-inferred subtype (S1-S3) versus HCs. **(B)** Subtype comparisons relative to NA. **(C)** Each ALS subtype compared to all other subtypes. **(D)** Pairwise subtype contrasts (S2 vs S1; S3 vs S1; S3 vs S2). For each panel, *t*-value maps display regions with significant volume differences or ventricular expansion: positive *t*-values (yellow-red) reflect expansion or higher volume in the first group, while negative *t*-values (blue) indicate lower volume. Light gray overlays flag regions where contrasts did not survive FDR correction (*p* > 0.05). Group differences were assessed using LME models. Sample sizes: HC = 144, S0 = 61, S1 = 65, S2 = 39, S3 = 33. ALS = amyotrophic lateral sclerosis; HCs = Healthy Controls; LME = Linear Mixed-Effects.

The Normal Appearing (S0) group showed no regional atrophy versus controls (i.e. no negative t-values) and instead displayed positive t-values in limbic and basal nuclei (e.g., hippocampus *t*=4.68, amygdala *t*=4.07, putamen *t*=3.92). Relative to all subtypes (Fig. 6C, left), S0 showed the greatest preservation across precentral, pallidal, accumbens, hippocampal, amygdalar, superior-temporal, putaminal, and superior-frontal regions, as well as the corpus callosum, consistent with a structurally intact, possibly compensatory, frontolimbic-striatal profile.

S1 exhibited the canonical motor/CST/brainstem atrophy pattern (e.g., corticospinal tract *t*=-9.69, brainstem *t*=-9.13, precentral *t*=-8.05) accompanied by ventricular enlargement (third *t*=5.18; lateral *t*=4.05). Compared with the S0 group (Fig. 6B, left), S1 showed extensive precentral and brainstem atrophy together with CST degeneration, consistent with early motor-system involvement. Relative to all other subtypes (Fig. 6C, left), S1 displayed relative sparing of the caudate, entorhinal cortex, and amygdala, indicating a selective motor-dominant trajectory with limited limbic participation.

S2 displayed a limbic-predominant pattern (amygdala *t*=-11.50, entorhinal *t*=-7.48, hippocampus *t*=-7.36), with a relatively spared postcentral gyrus (*t*=3.49) and no ventricular enlargement. In comparison to the S0 group (Fig. 6B, left), S2 showed pronounced limbic and medial-temporal atrophy but maintained intact motor and parietal cortices. Relative to other subtypes (Fig. 6C, left), S2 showed preservation in superior and rostral-middle frontal, superior-parietal, postcentral, precentral, paracentral, and hand-knob regions, consistent with a limbic-onset trajectory that later extends toward motor networks.

S3 showed a striatal plus fronto-parietal/motor profile (e.g., precentral *t*=-5.72, caudate *t*=-5.23, inferior parietal *t*=-4.89) with enlargement of the third and lateral ventricles. Compared with the S0 group (Fig. 6B, left), S3 demonstrated extensive fronto-striatal and parietal involvement (superior-frontal *t*=-4.16, rostral-middle-frontal *t*=-3.90, putamen *t*=-3.59, caudate *t*=-4.17) but relative preservation of the brainstem, cerebellum and CST. In the subtype-to-subtype contrasts (Fig. 6C, left), S3 exhibited relative preservation or enlargement of the brainstem, cerebellar white matter, ventral diencephalon, CST, and vermal lobules I-V and VIII-X, reflecting a fronto-striatal-dominant phenotype with maintained integrity of cerebellar-brainstem structures.

These separations were strongly validated by pairwise subtype contrasts (Fig. 6D, left): the S2 vs. S1 comparison highlighted the prominent limbic profile, while the S3 vs. S1 contrast emphasized the distinct striatal/fronto-parietal signature over the canonical motor pattern. Finally, the S3 vs. S2 contrast clearly demarcated the frontal-motor burden of S3 against the limbic atrophy characteristic of S2.

### Longitudinal contrasts revealed separable imaging signatures

The longitudinal analysis, which measures the rate of atrophy change over time, provided a dynamic view of disease progression, often revealing aggressive spread that was not yet the dominant feature in the cross-sectional burden. Note that no longitudinal information was used in deriving the subtypes.

Despite lacking baseline atrophy, the S0 group demonstrates clear progressive motor decline versus controls (Fig. 6A, right), with the fastest rates of tissue loss observed in the precentral Gyrus (*t*=-6.35), paracentral lobule (*t*=-6.31), and hand-knob (*t*=-5.38) regions. This suggests that while S0 is structurally intact at baseline, the pathology is already accelerating within the motor system. This atrophy is accompanied by a pronounced rate of ventricular enlargement (third ventricle *t*=4.86, fourth ventricle *t*=4.34 and lateral ventricle *t*=3.71). S0 differences compared to all subtypes did not survive FDR correction and are overlaid with light gray (Fig. 6C, right).

Compared to controls (Fig. 6A, right), S1 exhibited a high rate of atrophy progression concentrated in the primary motor system. The highest observed atrophy rate was in the precentral gyrus (*t*=-8.14). This atrophy was accompanied by further expansion in the lateral ventricle (*t*=5.57), inferior lateral ventricle (*t*=5.38), third ventricle (*t*=5.21), and fourth ventricle (*t*=3.45). Intriguingly, the caudate showed less atrophy progression than in healthy controls (*t*=3.56), indicating a relative sparing of this subcortical structure despite the aggressive motor pathway involvement. Other longitudinal S1 comparison contrasts did not survive FDR correction (Fig. 6BCD, right), except for the S3 vs S1 comparison (Fig. 6D, right), which is detailed below.

While cross-sectionally defined by limbic atrophy, S2 demonstrated a mixed longitudinal progression versus controls (Fig. 6A, right), with dominant motor-system decline (e.g., hand-knob *t*=-5.08, precentral gyrus (*t*=-4.48) and the brainstem (*t*=-4.77) accompanied by limbic atrophy. This atrophy was accompanied by a pronounced rate of ventricular enlargement (lateral ventricle *t*=5.33, third ventricle *t*=5.14, inferior lateral ventricle *t*=3.42, and fourth ventricle *t*=3.15). Other contrasts involving S2 did not survive multiple-comparison correction (light gray overlay in Fig. 6BCD, right).

S3 progression rate compared to controls (Fig. 6A, right) was dominated by caudal and descending motor structures atrophy. The highest rate of atrophy was observed in the brainstem (*t*=-6.75), followed by the ventral diencephalon (*t*=-5.85), CST (*t*=-5.26), anterior thalamic rad (*t*=-4.72), precentral gyrus (*t*=-4.63) and thalamus (*t*=-4.57). S3 showed significant differences that survived FDR correction when compared to other ALS groups. Specifically, S3 demonstrated a significantly faster progression of atrophy in the brainstem (*t*=-4.01), thalamus (*t*=-3.81), CST (*t*=-3.74), and anterior thalamic radiations (*t*=-3.67) compared to S0 (Fig. 6B, right). Furthermore, S3 showed a unique acceleration of CST atrophy compared to S1 alone (Fig. 6D, right) and when compared to all other subtypes combined (Fig. 6C, right). This robust finding confirms that S3 is defined by a uniquely aggressive rate of degeneration along the central motor and thalamic-frontal pathways. This atrophy was accompanied by a severe rate of expansion in the third ventricle (*t*=6.17) and lateral ventricle (*t*=4.96).

### Association Analysis: Clinical and Imaging Features with Disease Stages

Widespread associations between SusTaIn stage and regional atrophy (w-scores) were observed across subtypes (Fig. 7A). At the group level, combining all ALS cases (including S0), the strongest negative associations with disease stage, indicative of progressive tissue loss, were found in limbic and subcortical regions. The most significant atrophy was recorded in the amygdala (*t*=-10.45), thalamus (*t*=-9.82), ventral diencephalon (*t*=-9.07), accumbens area (*t*=-8.49), and then putamen (*t*=-8.37), followed by parahippocampal, precentral gyrus, and frontal regions. Conversely, ventricular volumes (lateral: *t*=6.76; third: *t*=6.74; inferior lateral: *t*=2.61) showed significant positive associations, reflecting the typical ventricular expansion that accompanies cerebral atrophy. Each subtype presented a distinct progression profile. S1 strongly supported a predominantly motor-subcortical trajectory, showing the tightest correlations between stage and atrophy in the brainstem (*t*=-6.82), ventral diencephalon (*t*=-6.12), precentral gyrus (*t*=-4.96), thalamus (*t*=-4.27), and the CST (*t*=-4.16). S2 exhibited robust associations in limbic-thalamic structures like the amygdala (*t*=-6.29), thalamus (*t*=-6.16), and accumbens (*t*=-6.09), but also involved the inferior fronto-occipital fasciculus (*t*=-4.77), superior frontal (*t*=-4.69), putamen (*t*=-4.66), and CST (*t*=-4.59), indicating that while limbic regions are primary, motor and thalamic structures become substantially involved later. S2 also showed significant ventricular expansion (lateral: *t*=5.19; third: *t*=3.75). Subtype 3 (S3) was characterized by the most extensive cortical-subcortical involvement, with significant atrophy in the amygdala (*t*=-5.20), inferior parietal (*t*=-4.62), ventral diencephalon (*t*=-4.24), anterior thalamic radiations (*t*=-4.09), and frontal regions (caudal/rostral middle frontal, superior frontal: *t*=-3.65). Like the S2, ventricular expansion was positively correlated with stage (third: *t*=5.18; lateral: *t*=3.54).

**Figure 7.**
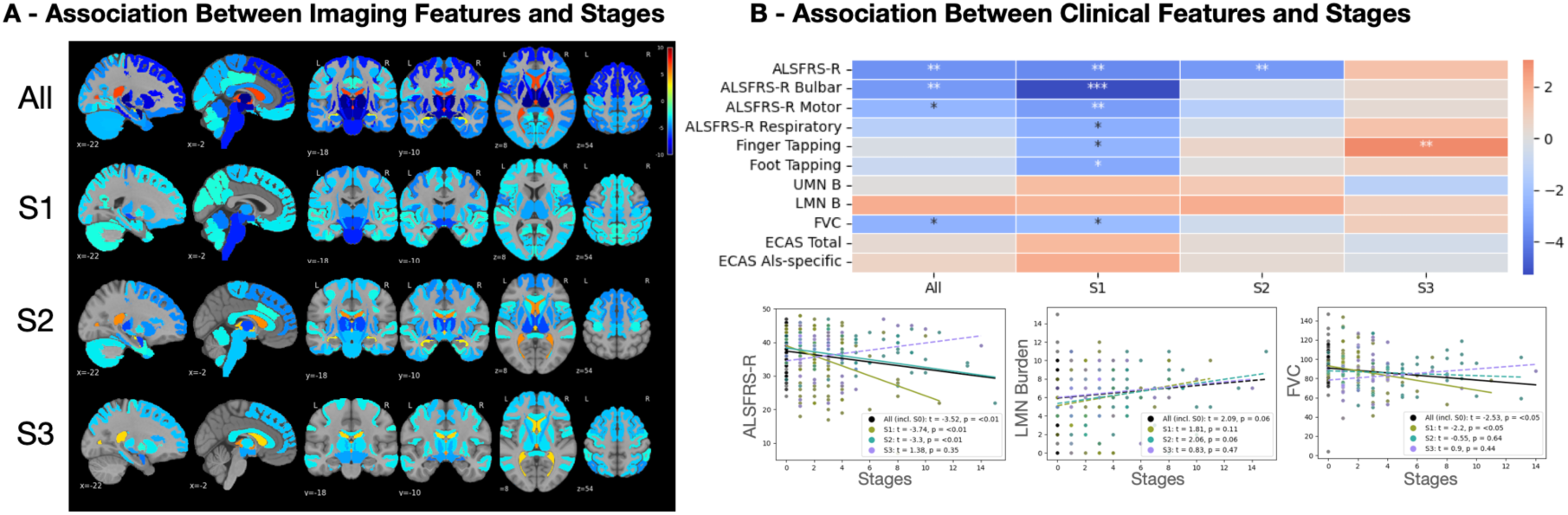
Associations between SuStaIn-inferred disease stage and longitudinal imaging or clinical features in ALS. **(A)** Brain maps display mixed-effects model associations (*t*-values, significant at *p* < 0.05 after FDR-correction) between SuStaIn stage and regional longitudinal imaging features (w-scores) for the overall ALS cohort including S0 (“All”) and by inferred subtype (S1-S3). Blue shades indicate negative associations (advanced stage = greater atrophy), while red hues indicate positive associations (advanced stage = relative expansion, most often ventricular). **(B) (Top)** Heatmap summarizing mixed-effects model associations (t-values, FDR-corrected) between SuStaIn stage and key clinical variables across subtypes. Negative values (blue) indicate functional decline with advancing stage, while for UMN/LMN burden, red shades denote that higher stage is related to increased motor burden. Asterisks indicate FDR-corrected statistical significance: * *p* < 0.05, ** *p* < 0.01, *** *p* < 0.001. **(Bottom)** Representative scatterplots show stage associations with ALSFRS-R total score, LMN burden, and FVC. Colored lines (dashed when *p* > 0.05) represent subtype-specific fits, with black lines indicating the combined cohort, including the S0 group. ALS = amyotrophic lateral sclerosis; ALSFRS-R = Amyotrophic Lateral Sclerosis Functional Rating Scale-Revised; FDR = False Discovery Rate; FVC = Forced Vital Capacity; LMN = Lower Motor Neuron; UMN = Upper Motor Neuron.

Across clinical measures (Fig. 7B), SuStaIn stage showed expected negative associations with functional decline. ALSFRS-R and FVC both decreased significantly with increasing stage when analyzed across all subtypes, with the strongest effects observed in the Motor/CST-Dominant (S1) subtype (*p* < 0.05). Motor performance, assessed by finger and foot tapping frequency, showed a significant stage-dependent decline in S1. However, the unexpected significant increase in finger tapping metrics seen in S3 at later stages is likely an artifact attributable to the limited number of subjects available for analysis at higher stages (Supplementary Fig. 1). LMN burden tended to increase with stage (p ≈ 0.06 after FDR), particularly in the combined and S2 groups, consistent with progressive lower motor neuron involvement. In contrast, UMN burden and ECAS scores showed no significant association. Representative scatterplots for ALSFRS-R, FVC, and LMN burden are shown in Fig. 7, with the full association map presented in Supplementary Fig. 1.

## Discussion

In the present study, we applied the SuStaIn algorithm to DBM-derived regional brain volumes from CALSNIC-1 and CALSNIC-2 cohorts to identify ALS subtypes. In addition to one “Normal Appearing/Motor-Onset” (S0) group, SuStaIn modeling of cross-sectional DBM brain imaging revealed three biologically plausible ALS trajectories that align with and extend our cross-sectional and longitudinal analyses. By combining a probabilistic disease progression model with longitudinal mixed-effects validation, this work provides evidence that ALS subtypes can be reproducibly staged and clinically interpreted. Subtype and stage consistency across visits (≥90%) confirmed the biological validity of the inferred trajectories and supports their potential for longitudinal patient stratification.

The SusTaIn-inferred Normal Appearing (S0) group and disease subtypes presented with unique imaging and clinical profiles:

**S0 (“Normal Appearing/Motor-Onset”):** appeared structurally intact at baseline, showing no cross-sectional atrophy relative to healthy controls and even relative hypertrophy in limbic and basal nuclei. Despite this preservation, longitudinal analysis revealed clear motor-system decline, accompanied by pronounced ventricular expansion, suggesting delayed but eventual motor involvement. This pattern may reflect early compensatory reorganization or neuronal dysfunction prior to manifest macrostructural atrophy, consistent with previous findings in ALS and related disorders.^44,45^ Clinically, individuals in S0 exhibited cognitive deficits, particularly in language and verbal fluency, suggesting early extra-motor pathology despite limited imaging atrophy.
**Subtype 1 (“Motor/CST-Dominant”):** exhibited the canonical ALS trajectory, beginning with corticospinal, precentral, paracentral, and brainstem atrophy, later spreading into frontoparietal and subcortical regions. These findings were supported by LME analyses showing prominent motor-system atrophy and progressive decline in motor and brainstem regions along with extension in ventricles compared to controls. When compared to S3, S1 has a lower progression of atrophy in CST. Stage-feature associations revealed strong stage dependence in the CST, thalamus, and brainstem, closely paralleling ALSFRS-R decline, confirming that SuStaIn staging captures biologically and clinically meaningful motor progression. Clinically, S1 presented greater UMN burden, lower ALSFRS-R scores, and intermediate survival (∼30 months), representing the classic ALS form.
**Subtype 2 (“Limbic-Onset with Motor Progression”):** followed a distinct trajectory characterized by early atrophy in limbic and medial temporal regions (amygdala, hippocampus, entorhinal cortex), later extending into motor and associative cortices. Cross-sectionally, S2 showed the most pronounced limbic atrophy; longitudinally, it evolved toward a mixed limbic-motor profile with ventricles extension. Longitudinal contrasts involving S2 did not survive multiple-comparison correction. Stage correlations confirmed this shift, showing limbic dominance cross-sectionally and emerging CST/paracentral associations longitudinally, consistent with a transition from limbic to motor involvement. Clinically, S2 featured balanced sex distribution, lower UMN burden, higher ALSFRS-R motor scores, a higher proportion of “Possible” diagnoses, and the lowest levels of cognitive dysfunction, consistent with a milder, slower-progressing phenotype despite intermediate survival (∼27 months).
**Subtype 3 (“Striatal-Fronto-parietal with Motor-Thalamic Rapid Progressor”):** displayed the most extensive and aggressive neurodegeneration. Early atrophy appeared in precentral and caudate regions, spreading to frontoparietal and striatal networks and later to limbic and motor-associative areas. Cross-sectional LME analysis confirmed dominant fronto-parietal and striatal atrophy with relative preservation of cerebellar white matter. Longitudinally, S3 was the only subtype to have a difference in progression of atrophy when compared to other subgroups. Specifically, S3 exhibited a markedly faster progression of atrophy in the brainstem, thalamus, CST, and anterior thalamic radiations when compared to S0, consistent with an aggressive motor-thalamic trajectory. This accelerated rate was further highlighted by the fact that CST atrophy in S3 had a significantly higher progression rate when compared to S1 alone and when compared to all other combined subgroups. Stage-feature associations showed that S3 progression was driven by amygdala, fronto-parietal, striatal, and thalamic correlations, supporting a frontal-subcortical network trajectory rather than isolated motor tract degeneration. Notably, CST and brainstem atrophy were time-dependent but not stage-dependent, suggesting rapid continuous decline decoupled from SuStaIn staging once ΔTime was controlled. This distinction highlights how integrating SuStaIn stage with longitudinal modeling differentiates true disease staging effects from simple temporal change. Clinically, S3 showed the most severe multidomain cognitive impairment (memory, language, fluency) and poorest survival (∼24 months).

Our findings align with and extend previous imaging-based ALS stratification efforts. Baumeister et *al.*,^46^ using CALSNIC and Utrecht data, applied the method of contrastive trajectory inference and identified three ALS subtrajectories: ALS_motor, ALS_diffuse, and ALS_limbic, based on gray matter density and white-matter fractional anisotropy. Their ALS_motor and ALS_limbic clusters closely correspond to our S1 (“Motor/CST-Dominant”) and S2 (“Limbic-Onset with Motor Progression”) subtypes, both defined by predominant motor and limbic degeneration, respectively. Our third subtype (S3) extends this framework by revealing extensive fronto-striatal and parietal atrophy at baseline and a uniquely aggressive rate of longitudinal degeneration along motor-thalamic pathways. Unlike Baumeister et *al.*,^46^ who analyzed cross-sectional data, we incorporated longitudinal validation, ΔTime-adjusted mixed-effects modeling to compare longitudinal change between groups and link SuStaIn-inferred stage with both clinical decline and DBM progression, distinguishing disease-stage effects from simple temporal change.

Our findings also extend the work of Shen et *al.*^24^ who applied SuStaIn to volumetric MRI data from ALS, ALS-FTD, and bvFTD cohorts, identifying one Normal Appearing group and two atrophy subtypes: Prefrontal/Somatomotor-predominant and Limbic-predominant. Their Prefrontal/Somatomotor subtype parallels our “Motor/CST-Dominant” (S1) trajectory, both showing early frontal and motor-cortical atrophy progressing to subcortical structures such as the thalamus and basal ganglia. Their Limbic-predominant subtype overlaps anatomically with our “Limbic-Onset with Motor Progression” (S2) trajectory, marked by early hippocampal, amygdalar atrophy; however, our S2 cases showed preserved cognition and higher ALSFRS-R motor scores and lower UMN burden, reflecting a limbic-onset ALS phenotype rather than an ALS-FTD continuum. In contrast to Shen et *al.*,^24^ our analysis was restricted to ALS-only participants, allowing us to isolate intrinsic motor and limbic heterogeneity within ALS rather than across the ALS-FTD spectrum. Moreover, our DBM-based approach incorporated longitudinal analysis between subtypes and time interval-adjusted mixed-effects modeling when linking SuStaIn-inferred stages to both clinical decline and imaging progression, confirming that inferred trajectories represent reproducible and biologically meaningful disease progression.

Compared with the probabilistic network-based clustering algorithm of Tan et *al.*^47^ who identified Pure Motor (PM), Fronto-Temporal (FT), and Cingulate-Parietal-Temporal (CPT) clusters using cortical thickness and white-matter measures, our SuStaIn-derived subtypes show both overlap and refinement. Our “Motor/CST-Dominant” (S1) subtype aligns closely with Tan’s PM cluster, both reflecting corticospinal, precentral, and brainstem degeneration with concurrent ventricular enlargement. The “Limbic-Onset with Motor Progression” (S2) shares partial limbic-temporal involvement with Tan’s FT subtype but remains restricted to ALS without overt frontotemporal cortical atrophy or cognitive impairment, suggesting a limbic-onset ALS phenotype distinct from ALS-FTD. Our “Striatal-Fronto-Parietal with Motor-Thalamic Rapid Progressor” (S3) shows the greatest convergence with Tan’s FT cluster, sharing extensive fronto-striatal, thalamic, and motor cortical degeneration, relative cerebellar sparing cross-sectionally, and most severe multidomain cognitive impairment. Overall, our results advance these prior frameworks by integrating temporal staging and longitudinal subtype comparison to distinguish dynamic disease trajectories from static structural clusters.

An advantage of SuStaIn over contrastive trajectory inference and probabilistic clustering approaches is that it explicitly models a Stage 0 (“normal-appearing”) group, representing individuals whose input features remain within the normal range of the inferred disease trajectories. This internally defined baseline enables interpretation of subtype- and stage-specific effects relative to a normative group derived directly from the disease progression model, a feature that is not available in models that derive only disease-associated trajectories or clusters without an explicit zero-stage subgroup.

Associations between SuStaIn stage and both imaging and clinical features confirmed the biological validity of the inferred trajectories. Stage correlated most strongly with atrophy in limbic-striatal and thalamic regions (amygdala, putamen, thalamus, ventral diencephalon), indicating that degeneration in these areas best reflects cumulative disease progression rather than simple aging effects associated with time. These stage-feature associations bridge cross-sectional and longitudinal perspectives: while SuStaIn infers progression from population-level heterogeneity, mixed-effects models capture within-person temporal change. Regions whose atrophy remained significantly correlated with stage after adjusting for ΔTime can thus be interpreted as genuine markers of disease advancement, distinguishing pathological staging from linear aging or follow-up duration. Parallel associations with clinical metrics reinforced the biological interpretability of SuStaIn staging. In the Motor/CST-Dominant subtype (S1), higher stages correlated with lower ALSFRS-R, FVC and tapping values and greater LMN burden, reflecting progressive decline in functional, respiratory, and motor-neuronal integrity. These relationships confirm that SuStaIn stage meaningfully indexes disease advancement along the motor axis. Paradoxical finger tapping associations with stage in S3 likely stem from sample imbalance and sparse representation at advanced stages rather than true inverse trajectories. Together, these findings indicate that SuStaIn stage captures the expected pattern of motor and respiratory deterioration across ALS progression, providing a biologically interpretable bridge between imaging-inferred stage and functional decline.

This study leveraged a fully data-driven SuStaIn approach applied to DBM-derived volumetric features in regions defined by Brettschneider’s neuropathological staging, yielding biologically interpretable subtypes aligned with known ALS mechanisms. Although based on cross-sectional imaging input, longitudinal validation of subtype stability and progression differences demonstrates temporal robustness and clinical relevance. Notably, this is the first longitudinal validation of a DBM-based SuStaIn model in ALS, providing both mechanistic and clinical evidence of its translational potential for patient stratification and trial enrichment.

Limitations include modest sample sizes within certain subgroups, particularly S2 and S3, which may reduce power to detect subtle differences. Furthermore, the relative hypertrophy observed in limbic and basal nuclei within the Normal Appearing group (S0) warrants further biological investigation to determine whether this reflects compensatory neural reorganization or methodological factors. Future studies should integrate genetic and molecular biomarkers to elucidate the biological pathways underlying subtype-specific atrophy patterns. Although this study leveraged a substantial multi-center longitudinal cohort of 198 ALS patients with follow-up assessments at 0, 4, and 8 months, further expansion to larger and longer-term datasets will be important to confirm the reproducibility of our findings and to more comprehensively characterize potential transitions across SuStaIn stages over time.

Collectively, these strengths, including the multi-cohort design, DBM-based sensitivity, longitudinal SuStaIn validation, neuropathologically grounded ROI selection, and explicit linkage between imaging-derived stages and clinical progression, advance ALS subtyping toward a dynamic precision-medicine framework for patient stratification and therapeutic targeting.

## Data Availability

All data produced in the present study are available upon reasonable request to the authors.

Imaging and clinical data can be requested from the CALSNIC consortium (https://calsnic.org/).

## Acknowledgements

We are grateful to the study participants, site PIs, research staff and MRI technologists. The authors also acknowledge use of Compute Canada (https://alliancecan.ca/en) resources for performing the image processing analyses in the presented work. We thank ChatGPT-5 for assistance with language refinement and sentence structure to enhance the manuscript’s overall clarity and readability. The authors remain fully responsible for the scientific content and conclusions.

## Funding

This project was supported by research funds from a ALS-Canada Brain Canada discovery grant and by a postdoctoral fellowship from ALS-Canada and Brain Canada Foundation. CALSNIC was funded by the Canadian Institutes of Health Research (CIHR), ALS Canada, Brain Canada and the Shelly Mrkonjic ALS Research Fund.

## Competing interests

The authors report no competing interests.

## Supplementary material

Supplementary material is available online.

---

**Appendix 1.**
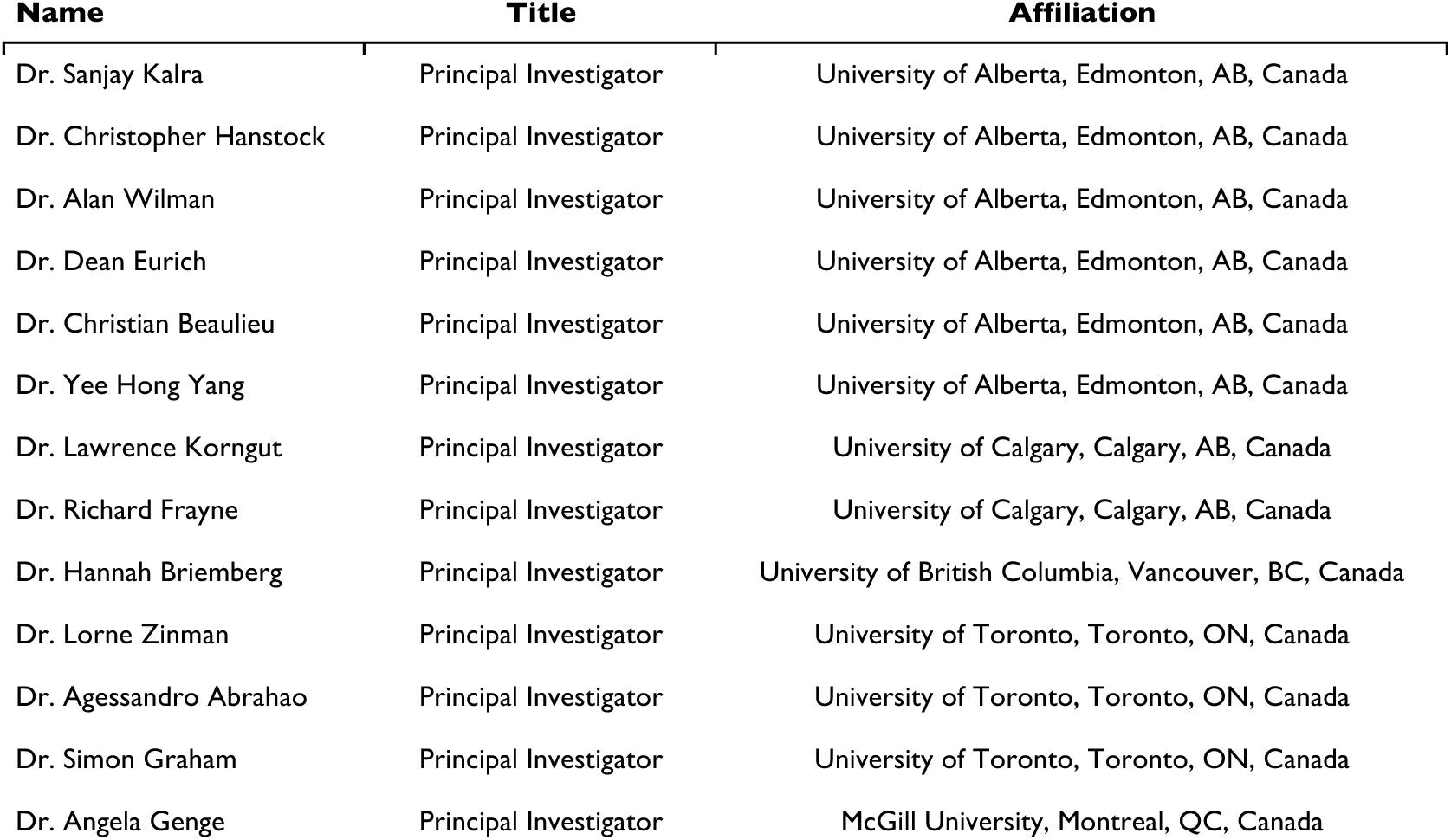
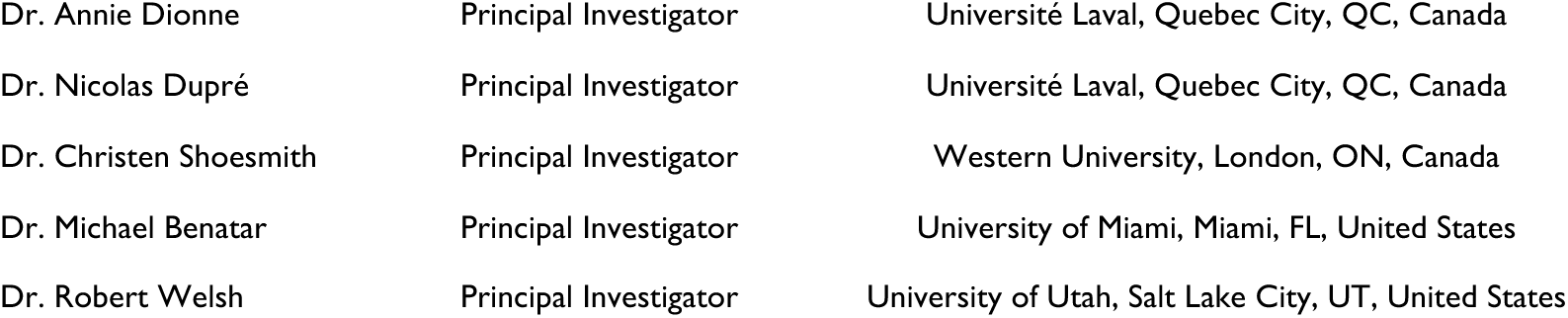
Membership of the Canadian ALS Neuroimaging Consortium (CALSNIC)

| **Name** | **Title** | **Affiliation** |
| --- | --- | --- |
| Dr. Sanjay Kalra | Principal Investigator | University of Alberta, Edmonton, AB, Canada |
| Dr. Christopher Hanstock | Principal Investigator | University of Alberta, Edmonton, AB, Canada |
| Dr. Alan Wilman | Principal Investigator | University of Alberta, Edmonton, AB, Canada |
| Dr. Dean Eurich | Principal Investigator | University of Alberta, Edmonton, AB, Canada |
| Dr. Christian Beaulieu | Principal Investigator | University of Alberta, Edmonton, AB, Canada |
| Dr. Yee Hong Yang | Principal Investigator | University of Alberta, Edmonton, AB, Canada |
| Dr. Lawrence Korngut | Principal Investigator | University of Calgary, Calgary, AB, Canada |
| Dr. Richard Frayne | Principal Investigator | University of Calgary, Calgary, AB, Canada |
| Dr. Hannah Briemberg | Principal Investigator | University of British Columbia, Vancouver, BC, Canada |
| Dr. Lorne Zinman | Principal Investigator | University of Toronto, Toronto, ON, Canada |
| Dr. Agessandro Abrahao | Principal Investigator | University of Toronto, Toronto, ON, Canada |
| Dr. Simon Graham | Principal Investigator | University of Toronto, Toronto, ON, Canada |
| Dr. Angela Genge | Principal Investigator | McGill University, Montreal, QC, Canada |
| Dr. Annie Dionne | Principal Investigator | Université Laval, Quebec City, QC, Canada |
| Dr. Nicolas Dupré | Principal Investigator | Université Laval, Quebec City, QC, Canada |
| Dr. Christen Shoesmith | Principal Investigator | Western University, London, ON, Canada |
| Dr. Michael Benatar | Principal Investigator | University of Miami, Miami, FL, United States |
| Dr. Robert Welsh | Principal Investigator | University of Utah, Salt Lake City, UT, United States |

## Notes

### Competing Interest Statement

The authors have declared no competing interest.

### Funding Statement

This study was supported by research funds from a ALS-Canada Brain Canada discovery grant and by a postdoctoral fellowship from ALS-Canada and Brain Canada Foundation. CALSNIC was funded by the Canadian Institutes of Health Research (CIHR), ALS Canada, Brain Canada and the Shelly Mrkonjic ALS Research Fund.

### Author Declarations

Ethics committee/IRB of University of Alberta Health Research Ethics Board (HREB) gave ethical approval for this work.

