## Supplementary for "Data-driven Subtyping and Staging of ALS: A Multicentre, Longitudinal, Deformation-Based Morphometry Study"

**Supplementary material**

A. Exclusion criteria

Patients aged below 40 years and those that received a diagnosis of other neurological or psychiatric conditions (e.g. bipolar disorder, brain trauma, epilepsy and depression) were excluded. Healthy control participants were excluded if they were aged below 40 years old or if they had history of cognitive impairment or neurological or psychiatric disorders. Participants who either did not undergo MRI scans or whose scans failed quality control assessments were also excluded.

B. Clinical evaluations

Clinical evaluation included the ALS Functional Rating Scale-Revised (ALSFRS-R),[28] from which the rate of disease progression rate (DPR) was estimated employing the formula: (48-ALSFRS-R)/symptom duration. Finger and foot tapping rates were calculated based on the average number of taps in 10 seconds over two independent trials from the left and right side. Left and right measurements were then averaged to obtain a unique score for finger and foot tapping. The Edinburgh Cognitive and Behavioural ALS Screen (ECAS), a multi-domain cognitive screening battery specifically developed for patients with ALS,[29] was also performed. Cognitive impairment was assessed using quantile regression-derived cutoffs for the North American version of the ECAS that accounts for age and education level[30] and transformed into percentage of abnormal values. As ECAS scores were collected longitudinally within the CALSNIC-2 study, but at baseline only within CALSNIC-1, this clinical feature was evaluated solely in a cross-sectional manner. A composite score was calculated to assess the degree of UMN involvement as indicated on neurological examination. This score integrates bilateral information from multiple clinical measures: spasticity (maximum of 4 points), muscle stretch reflexes (maximum of 8 points), the jaw jerk reflex (2 points), and presence of the Babinski sign (2 points), resulting in a total possible score of 16.[26] Similarly, a single LMN burden composite score was measured by considering hyporeflexia, muscle atrophy, and fasciculations across both the left (maximum score of 2 points each on both the right and left sides), as well as an additional point each for fasciculations in the face and tongue, and a point for tongue atrophy, culminating in a total possible score of 15. This composite score was designed to fully assess LMN burden across multiple spinal and bulbar segments. Additional clinical and demographic features of interest included site of (first symptom) onset, forced vital capacity (FVC), symptom duration at the first MRI visit, years of education, age and sex.

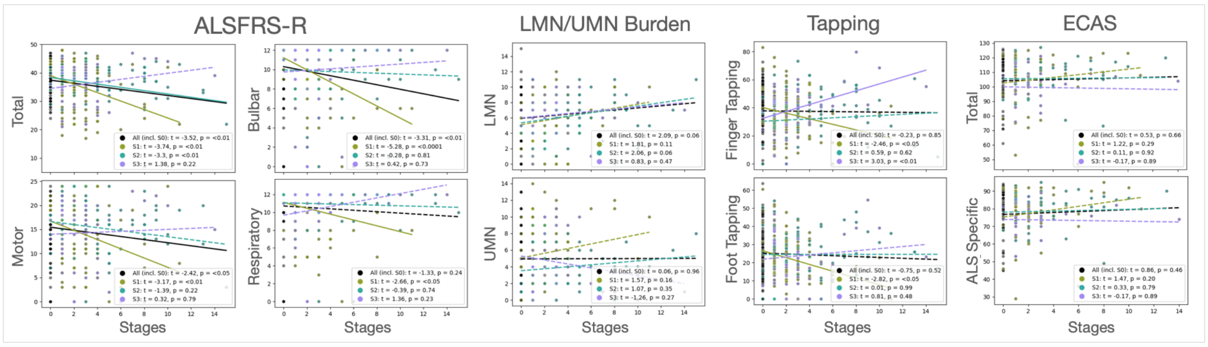


**Figure 1. Scatterplot visualization of SuStaIn stage-clinical feature associations.**

The scatterplots display observed longitudinal data (shared points) alongside model-predicted lines for each SuStaIn subtype (colored) and the overall patient cohort (black). Statistical associations (t and p values) were determined using linear mixed-effects models (adjusted for sex, age, and time since baseline) or ordinary least squares models for ECAS (adjusted for sex, age, and education years). Non-significant relationships ($p > 0.05$, FDR-corrected) are indicated by dashed lines and must be interpreted with caution. Note that the slopes at later disease stages, particularly within S3, are potentially biased due to the sparse data points.
